# Does Pecs II Block Reduce the Incidence of Post Mastectomy Pain Syndrome (PMPS)? A Cross Sectional Study

**DOI:** 10.1101/19006924

**Authors:** Vimal Varma, Chih N. Yeoh, Choon Y. Lee, Azrin M. Azidin

**Author notes:** **Address correspondence to:** Chih N. Yeoh, Department of Anesthesiology and Intensive Care, Universiti Kebangsaan Malaysia Medical Center, Jalan Yaacob Latif, Bandar Tun Razak, 56000 Kuala Lumpur, Malaysia. **Funding:** The authors have no sources of funding to declare for this manuscript.

## Abstract

**Background and Objectives:** Post mastectomy pain syndrome (PMPS) is a chronic pain condition that develops after breast cancer surgery. The objective of this study was to determine if Pecs II block administered prior to general anesthesia (GA) reduced the incidence of PMPS after mastectomy and axillary clearance (MAC) when compared with conventional analgesic therapy.

**Methods:** This cross sectional study compared incidence and severity of PMPS in 288 women who underwent unilateral MAC. Questionnaire survey was done via personal and telephone interview with 145 patients who received conventional analgesic therapy versus 143 patients who received Pecs II block. Outcomes assessed included incidence, severity and chronic pain symptom and sign score of PMPS.

**Results:** We found a significantly lower incidence of PMPS in patients who received Pecs II block compared with conventional analgesic therapy [49.7% vs. 63.4%, OR 0.57 (0.36-0.91), *P* = 0.018], which was a 22% relative risk reduction (RR 0.78). Severity of PMPS in Pecs II group was also significantly reduced as shown by lower static and dynamic pain scores at operative site (*P* < 0.001). Furthermore, Pecs II group reported significantly lower Chronic Pain Symptom and Sign Score (*P* = 0.002) compared to conventional group.

**Conclusions:** Pecs II block prior to MAC significantly reduced the incidence of PMPS, severity of chronic pain at operative site and number of chronic pain symptoms and signs related to PMPS.

This study is registered under National Medical Research Register. **NMRR ID: 17-2627-38056**

## INTRODUCTION

Breast cancer is the most common cancer in women and second most common cancer in the world comprising 25% of all newly diagnosed cancers in 2012.^1^ Breast cancer accounted for 32.1% of all cancer among females in Malaysia.^2^ It is the leading cause of cancer-related deaths among women. Surgery is still the mainstay of treatment despite therapeutic advances in management of breast malignancy over the last decade.^3^

Persistent pain following mastectomy was first reported in 1978.^4^ It is a distinctive, persistent and debilitating neuropathic pain syndrome and has been named post mastectomy pain syndrome (PMPS).^5^ It is a difficult clinical condition to treat, may have profound negative impact on health-related quality of life, and produce considerable disability and psychological distress.^6^ The incidence of PMPS is reported to range between 11% and 60%.^7-10^

The exact etiology of PMPS is not clear but studies have found positive correlations between the intensity of acute postsurgical pain and the development of chronic pain post breast cancer surgeries.^11-13^ Over the years, several regional anesthesia techniques such as thoracic epidural, paravertebral block, intercostal nerve block and intra-pleural block was being utilized in breast surgeries to optimize acute post-operative pain.^14^

The advancement of ultrasound (US) technology in the field of regional anesthesia has led to the introduction of a lesser invasive block, pectoral nerves block (Pecs block, later known as Pecs I block) by Blanco in 2011.^15^ It is a superficial inter-fascial block involving placement of local anesthetic (LA) between pectoralis major and minor muscles in order to anesthetize the lateral and medial pectoral nerves. A modified Pecs block (Pecs II block) was introduced by the same author a year later.^16^ Additional LA injection into the inter-fascial plane between the pectoralis minor and serratus anterior muscles anesthetizes the anterolateral chest wall by blocking the intercostobrachial, third to sixth intercostal and long thoracic nerves. Due to its wider coverage, Pecs II block has gained popularity and is now the block technique of choice in breast surgeries.^17,18^

To the best of our knowledge, there are no published studies on the effectiveness of Pecs II block in reducing the incidence and/or severity of PMPS. By conducting a questionnaire survey on patients who had undergone mastectomy and axillary clearance (MAC) in our institution, we hoped to be able to shed some light on the relationship between Pecs II block and PMPS.

## METHODS

This cross sectional study was approved by the research and ethics committees in both institutional and national levels. This project was registered under National Medical Research Register (NMRR), Ministry of Health Malaysia.

Patients who underwent MAC from July 2015 – June 2017 in Hospital Kuala Lumpur (HKL) were recruited. In order to standardize surgical technique and reduce confounding factors, case selection was only limited to unilateral MAC. Other breast cancer surgeries, such as wide local excision, modified radical mastectomy (MRM) and lumpectomy, were excluded. Patients with the following characteristics and problems were excluded from our study – past history of chronic pain and on regular analgesics, chemotherapy or radiotherapy before surgery, surgical complications (such as infection or wound breakdown) or cancer recurrence, history of psychiatric illness, inability to be contacted, inability or unwillingness to participate in the study.

There is currently no standard definition for this chronic pain syndrome. Depending on the definitions applied, patient selection and methods used, the incidence varies. The definition of PMPS used in this study was modified from International Association for the Study of Pain^19^ (IASP) and other studies.^9,20^ In our study, PMPS is taken as chronic post surgical pain in the anterior aspect of the thorax, axilla, and/or upper half of the arm beginning after mastectomy, without objective evidence of local abnormality, persisting either continuously or intermittently for more than three months after surgery, and may be associated with allodynia or sensory loss.

Questionnaire used in this study was obtained with permission from Dr Manoj Kumar Karmakar, the author of a previous study on the effect of thoracic paravertebral block (TPVB) on chronic pain post mastectomy.^21^ It was prepared in both English and Malay versions **(Appendix 1)** to cater for our local population. The English version was translated to Malay using forward and retrograde translation methods to check the reliability and precision of the words by four independent translators who were bicultural Malay speakers with a good command of English. The translation was further evaluated by a panel of experts in the field of Regional Anesthesia and Pain Management to verify the idiomatic and cross-cultural equivalence to the English version. Seven respondents were then recruited to assess face validity of the questionnaire. Repeat evaluation was done until respondents understood all the questions in the process of content validation. No modification was needed as the first draft of questionnaire was well understood by all respondents. Internal consistency reliability and construct validation was done using Cronbach’s alpha, Kaiser-Meyer-Olkin (KMO) test and Bartlett’s test of sphericity.^22^ The KMO value of 0.56 and the significant Bartlett’s test of sphericity (P< 0.001) in this validation was acceptable. Cronbach’s alpha was 0.736, indicating good internal consistency.

Personal details, medical history and contact number of all patients who underwent MAC within the specified period were collected from patient’s medical records with the permission from the Head of Breast and Endocrine Unit, Department of General Surgery, HKL. Each patient who fulfilled the selection criteria was assigned a study number and grouped into either conventional analgesic therapy (Group A) or Pecs II block (Group B).

All patients included in this study had undergone MAC under GA with regular method of induction and maintenance inhalational anesthesia. Those who received Pecs II block had the procedure done before induction of anesthesia by a team of experienced regional anesthesiologists. Both groups received appropriate doses of opioid and nonsteroidal anti-inflammatory drugs (NSAID) for intraoperative analgesia. Post operatively, all patients received standardized oral analgesics for pain control.

The survey was conducted by a single investigator. Recruited patients were seen in the surgical outpatient clinic to obtain consent and complete the questionnaire. Patients who were not seen in the clinic were contacted by telephone to obtain verbal consent and answer the questionnaire according to the template prepared **(Appendix 2)**. A series of questions related to chronic pain of PMPS were asked as per questionnaire.

PMPS was deemed to be present in patients who reported pain at rest over the operated site, axilla, or arm (i.e., reported yes to any of the 3 questions 4, 6, and 8 in **Appendix 1**) related to surgery. Pain scores were recorded according to numerical rating scale (NRS), where 0 = no pain and 10 = worst imaginable pain. Pain scores of 1-3, 4-6 and 7-10 were graded as mild, moderate and severe pain respectively as per WHO Pain Ladder.^23^ Also in order to quantify and statistically compare the total number of chronic pain symptoms and signs reported by the patients, a score of 1 was given to a response “yes,” and a score of 0 was given to a response “no” to the questions listed in **Appendix 3**.^21^ Maximum possible score was 12. Patients suffering from PMPS were offered further assessment and management at our Pain Clinic.

### Statistical Analysis

Using Krejcie and Morgan formula,^24^ sample size was calculated based on the incidence of PMPS, which was our primary outcome variable. Published data showed that the incidence of PMPS ranges from 11% to 60%.^7-10^ With expected incidence of 60%, using 2 proportions sample test, we calculated that a minimum of 95 patients per study group would provide 80% power (α = 0.05).

Data were analyzed using SPSS version 23.0 software. Continuous variables are presented as mean ± standard deviations or median (interquartile range) where appropriate. Categorical data is shown as numbers and percentages. Comparison of continuous data between groups was performed by Student’s *t* test. The Chi square test was used to compare groups with categorical variables. A *P* value ≤ 0.05 was considered statistically significant. The odds ratio (OR) and relative risk or risk ratio (RR), its standard error and 95% confidence interval (CI) were calculated according to Altman.^25^

## RESULTS

A total of 288 out of 432 patients who underwent MAC in HKL during our study period were recruited. As shown in **Table 1**, there were no significant differences in terms of age and ethnicity between the two groups.

**TABLE 1.** Age and Ethnicity of Patients. Values expressed as mean ± standard deviation or frequency (percentage), where appropriate.

|  | **Group A** | **Group B** | **Overall** | ***P*** |
| --- | --- | --- | --- | --- |
| **Age** |  |  |  |  |
| | 55.9±11.0 | 56.2±9.8 | 56.1±10.4 | 0.991 |
| **Race** |  |  |  |  |
| Malay | 76 (52.4) | 88 (61.5) | 164 (56.9) | 0.068 |
| Chinese | 39 (26.9) | 19 (13.2) | 58 (20.1) | 0.581 |
| Indian | 25 (17.2) | 35 (24.4) | 60 (20.8) | 0.309 |
| Others | 5 (3.4) | 1 (0.7) | 6 (2.1%) | 0.546 |

A total of 163 patients (92 from Group A and 71 from Group B) reported postoperative pain consistent with the definition of PMPS applied in this study, giving an overall incidence of 56.6% **(Figure 2)**. There was a significantly lower incidence of PMPS in Group B compared to Group A (*P* = 0.018). Odds ratio for PMPS in Group B was 0.57 (CI 0.35-0.91) while relative risk was 0.78 (CI 0.64-0.96).

The mean chronic pain score on a scale of 0 to 10 was relatively low in both groups (**Table 2**). In terms of severity of PMPS, lower static and dynamic pain scores at operative site (*P* < 0.001) was noted in Group B. There was no significant difference found in pain severity at ipsilateral axilla and arm. No patient from both groups reported severe pain at operative site, ipsilateral axilla or arm.

**Table 2:** Chronic Pain Score based on WHO Pain Ladder. Data are presented as mean±SD.

|  | **Group A** | **Group B** | ***P*** |
| --- | --- | --- | --- |
| Pain at operative site at rest | 0.99±1.40 | 0.29±0.69 | < 0.001 |
| Pain at operative site upon arm movement | 1.41±1.92 | 0.42±0.88 | < 0.001 |
| Pain at ipsilateral axilla | 0.71±1.24 | 0.5±0.99 | 0.278 |
| Pain at ipsilateral arm | 0.54±1.12 | 0.37±0.82 | 0.333 |

**FIGURE 1.**
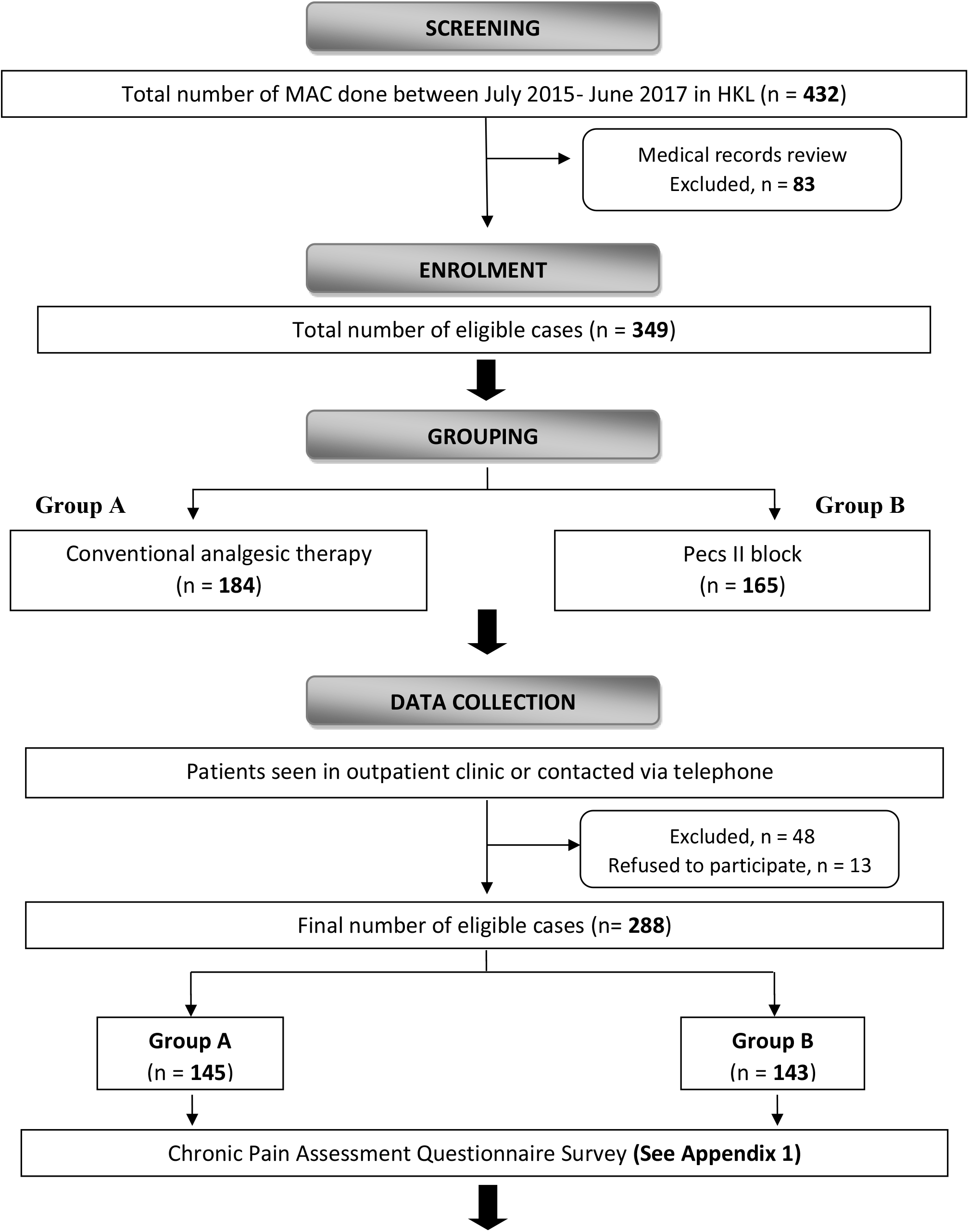
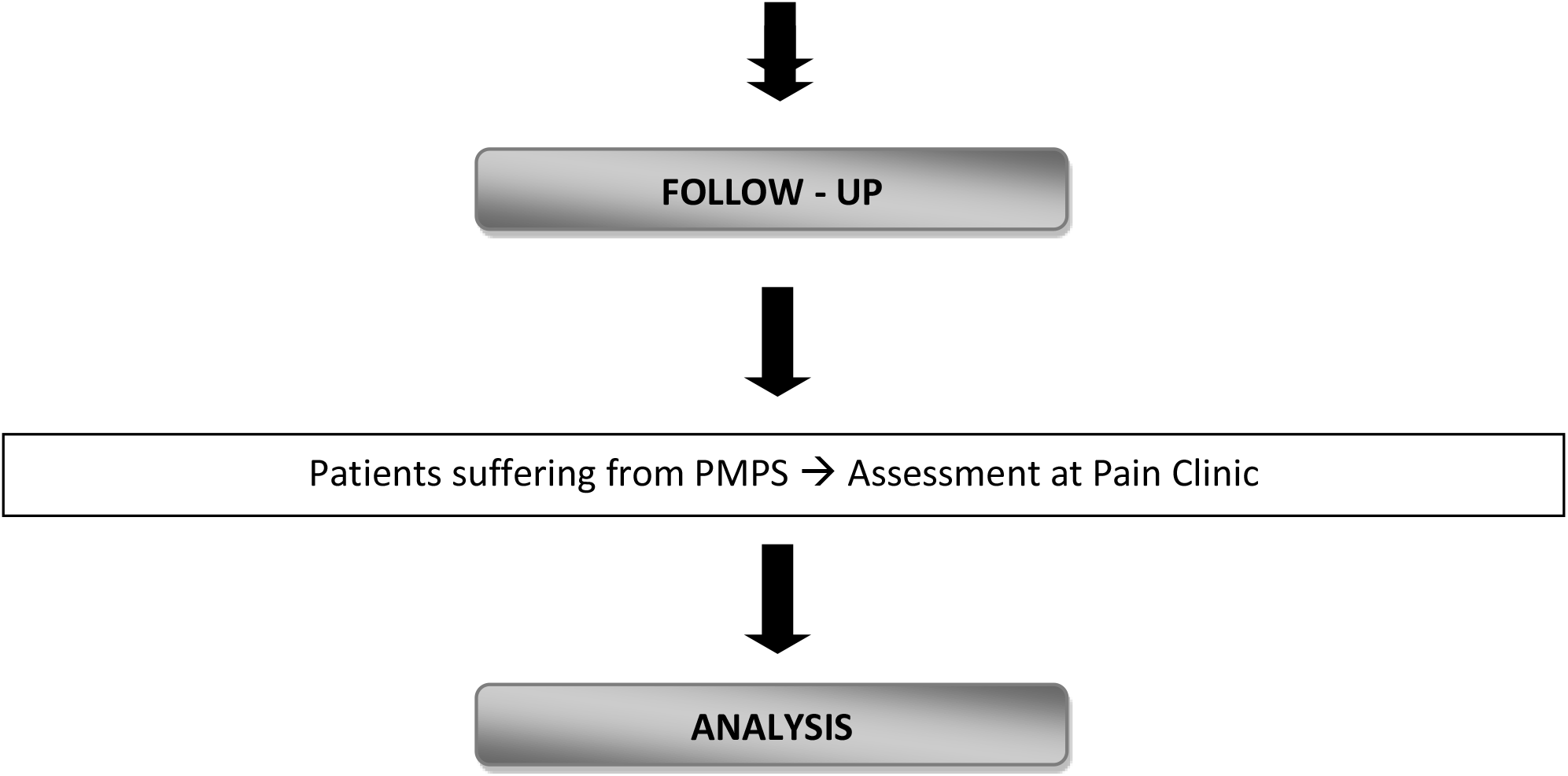
Flowchart.

**FIGURE 2.**
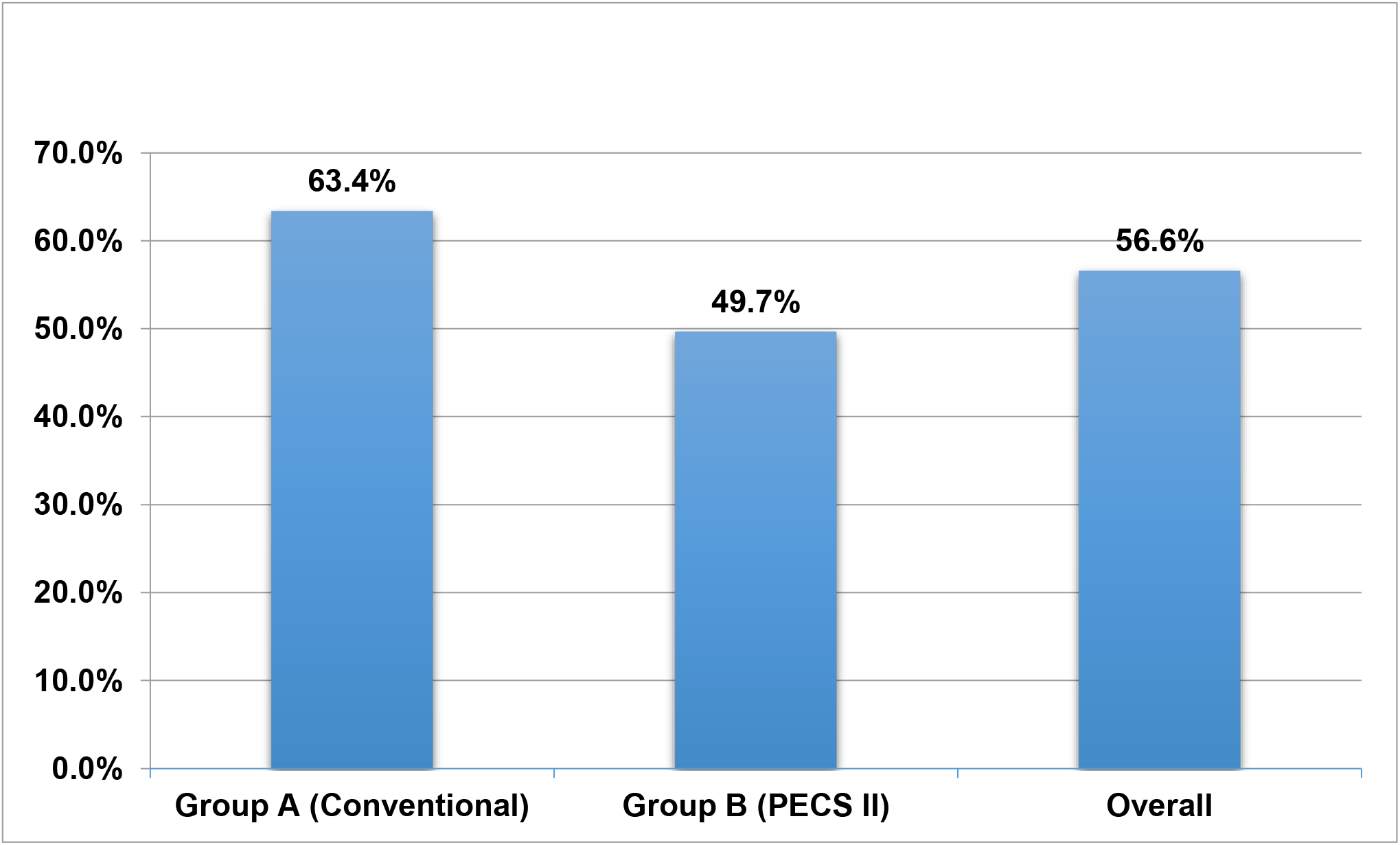
Incidence of PMPS.

The Chronic Pain Symptoms and Signs Score was significantly lower in Group B compared to Group A (1.91±1.41 vs. 1.43±1.18, *P* = 0.002). Pecs II group showed significantly lesser use of analgesics (P = 0.002) and low incidence of allodynia (P < 0.001). Almost similar percentage of patients reported painful phantom breast sensation in both groups (11.7 in Group A and 12.6 from Group B, *P* = 0.823). There was no significant difference noted in absent or reduced sensation at chest wall, axilla and arm in both groups.

## DISCUSSION

The overall incidence of PMPS of 56.6% in our study is similar to those reported in recent studies.^7-10^ Our study showed that incidence and severity of PMPS are significantly lower in patients who received Pecs II block for MAC. Odds ratio value of 0.57 (OR<1) indicates that the odds of developing PMPS in Pecs II group is 43% less likely than conventional analgesic therapy group with the true population effect between 64% and 9%. In other words, conventional group is 1.76 times more likely to develop PMPS. This result is statistically significant as the 95% CI (0.36-0.91) does not include the value of 1. However, since the upper limit CI of 0.91 is near to the value of 1, the practical significance of this finding is questionable. Also, OR can substantially underestimate (if OR<1) the magnitude of risk.^26^

Hence, we calculated the relative risk or risk ratio (RR). The RR value of 0.78 with 95% CI of less than 1 (0.64 – 0.96) once again proves that there is statistically significant evidence that Pecs II group is at lower risk of developing PMPS. The relative risk reduction (RRR) of 22% in developing PMPS is seen in Pecs II group compared to conventional analgesic therapy group. And again, since the upper limit CI of 0.96 narrowly misses the value of 1, the clinical significance of this finding is questionable.

Post-mastectomy pain syndrome (PMPS) is a form of chronic post surgical pain (CPSP) after breast cancer surgery that is characterized by pain and sensory disturbances consistent with a neuropathic origin.^7,8^ There is no accepted laboratory or radiographic criteria used to diagnose PMPS; it is a diagnosis of exclusion. The exact pathophysiology of PMPS is uncertain. It could be due to nerve damage, particularly the intercostobrachial nerve, as a result of excessive mechanical forces (severance, compression, ischemia, stretching or retraction) during operative procedures.^9^ The development of PMPS may also be related to molecular events that cause sensitization of peripheral nerves as well as spinal and supra-spinal processing centers.^27^

Duration and severity of acute postoperative pain is an important predictor of the development of CPSP^10^ and several studies showed association between acute postsurgical pain and development of chronic pain post breast surgeries.^11-13^ Hence, an effective acute pain management with peripheral nerve block (PNB) prior to surgery plays a significant role in reducing the risk of peripheral and central sensitization, and thereby mitigating development of PMPS.^27,28^ This was reflected in our study, in which the incidence and severity of PMPS was significantly lower in patients who received Pecs II block prior to surgery.

Over the past decade, multiple studies have been done to evaluate the role of PNB for preemptive as well as preventive analgesic therapy to improve acute post-operative pain and CPSP. Recent meta-analysis by Hussain et al^29^ and Terkawi et al^30^ provided moderate quality evidence suggesting that thoracic paravertebral block (TPVB) may potentially reduce the incidence of CPSP after breast surgery. However, TPVB is technically more challenging and poses higher risk of complications such as pneumothorax, spinal cord trauma, sympathetic block, and hypotension.^31^

Ultrasound-guided Pecs II block has gained popularity in breast cancer surgeries due to its relative simplicity, safety and efficacy.^17^ Studies by Wahba and Kamal^32^ as well as Kulhari et al^33^ reported significantly prolonged duration of postoperative analgesia with less requirement of rescue analgesia after breast cancer surgery in patients receiving a Pecs II block compared with a thoracic paravertebral block during the first 24 hours postoperative period. Recent systematic review and meta-analysis by Versyck et al^17^ and Singh et al^18^ also showed Pecs II block significantly alleviate acute postoperative pain and reduced opioid consumption intraoperatively as well as 24 hours post surgery. This opioid sparing effect is crucial in prevention of PMPS as opioid induced hyperalgesia is a risk factor for development of chronic post surgical pain.^28^ Thus Pecs II block has been recommended as first line option for regional analgesia in breast surgery. However, investigation into effectiveness of Pecs II block in these studies was only confined to the acute postoperative period, and none of the above studies addressed the issue of CPSP or PMPS.

There were a few limitations in our study. One limitation was the possibility of recall bias on pain memory among our respondents. Unfortunately, studies on this subject have been inconclusive. Salovey et al^34^ investigated the accuracy on reporting chronic pain episodes during health surveys and concluded that retrospective self-reports of pain collected systematically with measures of proven reliability seemed relatively trustworthy. However, in a later publication, Miranda et al^35^ concluded that prior musculoskeletal symptoms were poorly remembered after some years, and the recall was strongly influenced by current symptoms. Since the patients in our study underwent MAC not more than 2 years ago, we hope that recall bias was not a significant problem. Secondly, data on intra- and postoperative pain management were not collected in our study. These data could be relevant, given that ineffective pain control during the acute period could impact upon subsequent development of CPSP.

Another limitation was the use of postoperative adjuvant chemo- and/or radiotherapy in some patients. It is possible that such adjuvant therapy given in the perioperative period may lead to the development of PMPS by inducing local subclinical necrosis, neuritis, and myositis/fibrosis.^5,9,11^ In our study, only patients who received adjuvant therapy preoperatively, but not those in the postoperative period, were excluded. Though postoperative chemo- and/or radiotherapy did not significantly affect the incidence and severity of PMPS in our study, this finding is limited due to the variation in adjuvant therapy received based on disease condition.

### Conclusion

Pecs II block prior to MAC significantly reduced the incidence of PMPS, severity of chronic pain at operative site and number of chronic pain symptoms and signs related to PMPS.

## Data Availability

available at dropbox

https://www.dropbox.com/sh/atd3k409gjs79bn/AABBUulP19r7wEREIihHSRr2a?dl=0

## ACKNOWLEDGEMENTS

The authors thank Dr Manoj Kumar Karmakar, for his kind permission to use the chronic pain assessment questionnaire in our study. Special thanks to all the staffs of surgical department and record unit in HKL for their assistance during the conduct of this study. We wish to acknowledge the co-operation from all the patients who participated. We also would like to thank the Director General of Health Malaysia for his permission to publish this article.

## Appendix 1 QUESTIONNAIRE (Chronic Pain Assessment)

Study Number : ___________________ Contact No: ___________________

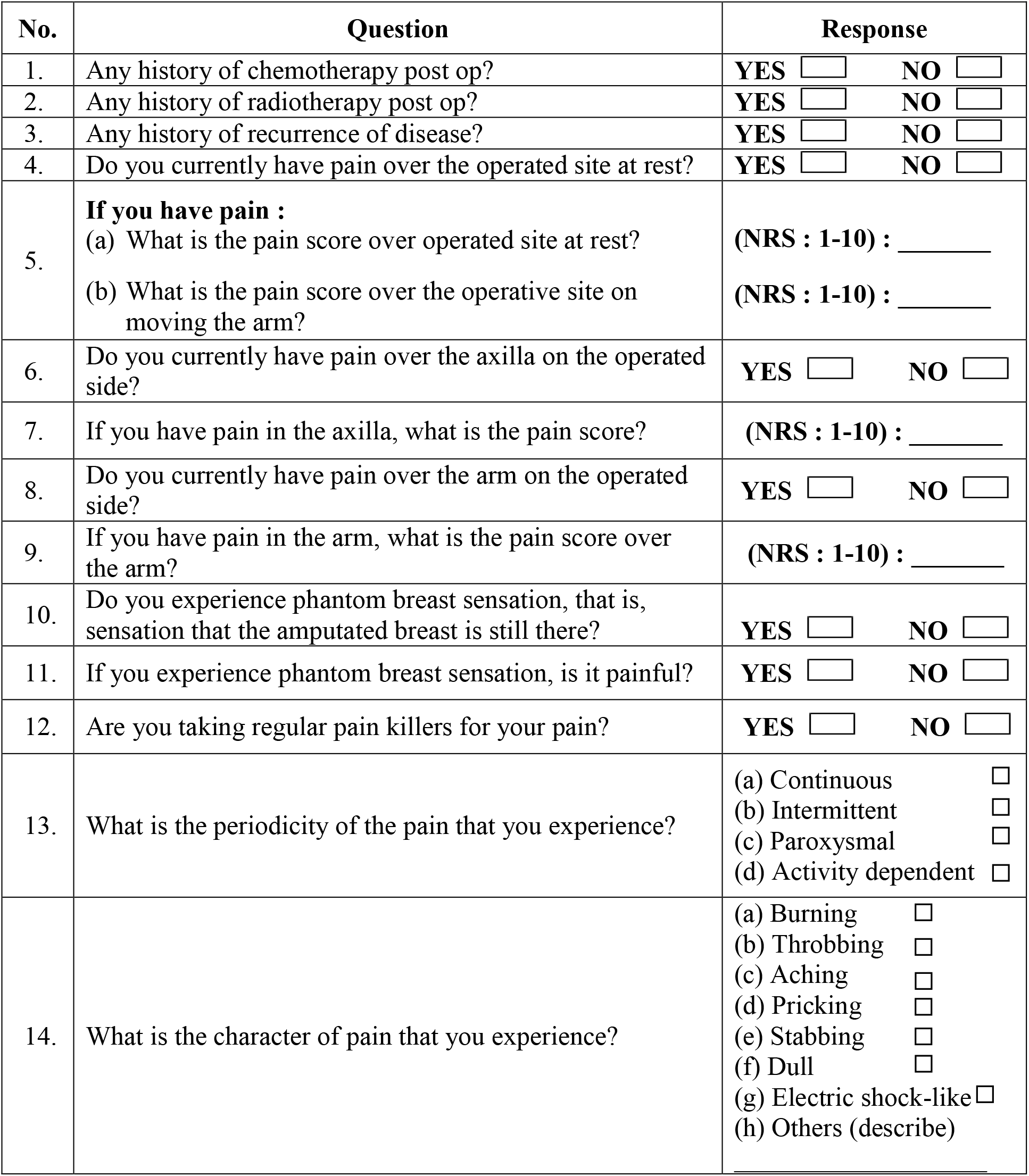
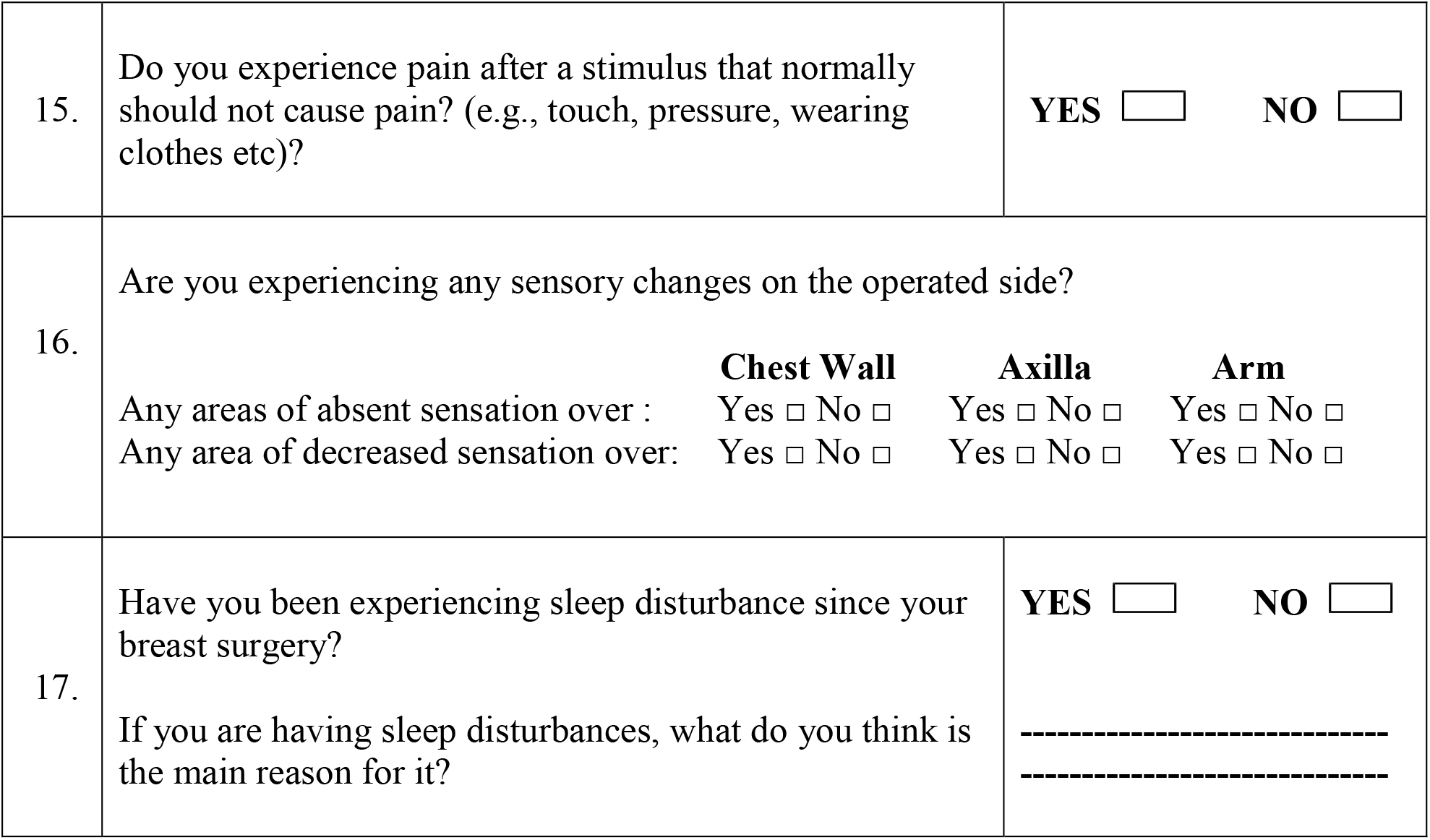

## Appendix 2 TELEPHONE INTERVIEW TEMPLATE

### Introduction

Hello Ms. / Mrs. ___________. Good morning/afternoon/evening.

I am **Dr Vimal Varma** from Department of Anesthesia and Critical Care, Hospital Kuala Lumpur. I’m currently conducting a survey related to the anesthetic service you received for your breast surgery done on [dd/mm/yyyy] in HKL.

### Purpose

This phone call is to find out how you are after the breast surgery. In particular, we would like to know whether you have problem with persistent pain over the area you are operated on, your armpit or your arm. This condition is called post mastectomy pain syndrome or PMPS. I will be asking you a few questions and the whole survey will take about **10 minutes**.

### Additional information for patients who have received Pecs II block

You may remember that when you had an anesthetic for the surgery, you have received an injection called pectoral nerve block for pain relief. The purpose of this survey is to determine whether that injection has any effect in reducing the occurrence of PMPS.

### Reassurance

Your participation in this study is entirely voluntary. The information collected from you will remain strictly confidential. You do not have to give reasons if you prefer not to take part, and your decision will not affect your future treatment. When you participate in this study, you do not have to pay anything; similarly, no payment will be given to you.

As I have mentioned, this survey will take about **10 minutes**, and your participation is greatly appreciated. If you are not free at the moment, we can reschedule this call when you are convenient to talk.

Do I have your kind permission to begin the survey?

### Questionnaire

Questions 1-17

### End the call with

These are all of the questions I have for you. Thank you so much for your time.

You will receive a written consent form and a patient information sheet (PIS) by mail in near future. Kindly retain the PIS, sign the consent form and mail the signed consent form in an enclosed stamped envelope to us.

If you have questions about this survey, you can contact me at this number 0123070598 for more information.

Thank you again and have a nice day.

**\*\* Remarks:**

If the patient suffers from PMPS, she will be offered further assessment and management at our Pain Clinic.

## Appendix 3 Chronic Pain Symptoms and Signs Score

(Maximum Possible Score : 12)

Study Number : ___________________ Contact No: ___________________

| No. | Question | Response |
| --- | --- | --- |
| 1 | Do you currently have pain over the operated site at rest? | YES ☐ NO ☐ |
| 2 | Do you currently have pain over the axilla on the operated side? | YES ☐ NO ☐ |
| 3 | Do you currently have pain over the arm on the operated side? | YES ☐ NO ☐ |
| 4 | If you experience phantom breast sensation, is it painful? | YES ☐ NO ☐ |
| 5 | Are you taking regular pain killers for your pain? | YES ☐ NO ☐ |
| 6 | Do you experience pain after a stimulus that normally should not cause pain? (e.g., touch, pressure, wearing clothes etc)? | YES ☐ NO ☐ |
| 7-12 | Any areas of absent sensation over : |  |
|  | 7. **Chest Wall** | Yes ☐ No ☐ |
|  | 8. **Axilla** | Yes ☐ No ☐ |
|  | 9. **Arm** | Yes ☐ No ☐ |
|  | Any areas of absent sensation over : |  |
|  | 10. **Chest Wall** | Yes ☐ No ☐ |
|  | 11. **Axilla** | Yes ☐ No ☐ |
|  | 12. **Arm** | Yes ☐ No ☐ |

## REFERENCES

1. Ferlay J, Soerjomataram I, Dikshit R, et al. Cancer incidence and mortality worldwide: sources, methods and major patterns in GLOBOCAN 2012. Int J Cancer. 2015;136:E359–E386.

2. Ministry of Health Malaysia. National cancer registry report: Malaysia cancer statistics-data and figure 2007-2011. http://www.care.upm.edu.my/dokumen/13603_NCR2007.pdf. Published June, 2015. Accessed November, 2017.

3. El-Tamer MB, Ward BM, Schifftner T, Neumayer L, Khuri S, Henderson W. Morbidity and mortality following breast cancer surgery in women: national benchmarks for standards of care. Ann Surg. 2007;245(5):665–671.

4. Wood, KM. Intercostobrachial nerve entrapment syndrome. South Med J. 1978;71:662–663.

5. Jung BF, Ahrendt GM, Oaklander AL, et al. Neuropathic pain following breast cancer surgery: proposed classification and research update. Pain. 2003;104:1–13.

6. Tasmuth T, Estlanderb AM, Kalso E. Effect of present pain and mood on the memory of past postoperative pain in women treated surgically for breast cancer. Pain. 1996;68:343–347.

7. Vilholm OJ, Cold S, Rasmussen L, et al. The post mastectomy pain syndrome: an epidemiological study on the prevalence of chronic pain after surgery for breast cancer. Br J Cancer. 2008;99:604–610.

8. Gartner R, Jensen MB, Nielsen J, et al. Prevalence of and factors associated with persistent pain following breast cancer surgery. J Am Med Assoc. 2009;302(18):1985–1992.

9. Smith WCS, Bourne D, Squair J, Phillips DO. A retrospective cohort study of post mastectomy pain syndrome. Pain. 1999;83(1):91–95.

10. Ballantyne JC, Cousins MJ, Giamberardino MA, et al. Chronic pain after surgery or injury. Pain. 2011;XIX(1):1–5.

11. Tasmuth T, Kataja M, Blomqvist C, Smitten KV, Kalso E. Treatment-related factors predisposing to chronic pain in patients with breast cancer. Acta Oncol. 1997;36(6):625–630.

12. Geving K, Molter H, Elisabeth H, Kroman N, Kehlet H. Predictive factors for the development of persistent pain after breast cancer surgery. Pain. 2015;156(12):2413–2422.

13. Poleshuck EL, Katz J, Andrus CH, et al. Risk factors for chronic pain following breast cancer surgery: A prospective study. J Pain. 2006;7(9):626–634.

14. Westbrook AJ, Buggy DJ. Anaesthesia for breast surgery. BJA CEPD Rev. 2012;3(5):151–154.

15. Blanco R. The ‘Pecs block’: a novel technique for providing analgesia after breast surgery. Anesthesia. 2011;66:847–848.

16. Blanco R, Fajardo M, Maldonado TP. Ultrasound description of Pecs II (modified Pecs I): A novel approach to breast surgery. Rev Esp Anestesiol Reanim. 2012;59:470–475.

17. Versyck B, Geffen G Van, Chin K. Analgesic efficacy of the Pecs II block: a systematic review and meta-analysis. Anaesthesia. 2019;74:663–673.

18. Singh PM, Borle A, Kaur M, Trikha A, Sinha A. Opioid-sparing effects of the thoracic interfascial plane blocks: A meta-analysis of randomized controlled trials. Saudi J Anaesth. 2018;12:103–111.

19. International Association for the Study of Pain. Classification of chronic pain. Descriptions of chronic pain syndromes and definitions of pain terms. Pain. 1986;3:142–143.

20. Waltho D, Rockwell G. Post-breast surgery pain syndrome: establishing a consensus for the definition of post-mastectomy pain syndrome to provide a standardized clinical and research approach — a review of the literature and discussion. Can J Surg. 2016;59(5):342–350.

21. Karmakar MK, Samy W, Li JW, et al. Thoracic paravertebral block and its effects on chronic pain and health-related quality of life after modified radical mastectomy. Reg Anesth Pain Med. 2014;39:289–298.

22. Hutcheson G, Sofroniou N. Introductory statistics using generalized linear models. In: The Multivariate Social Scientist. Thousand Oaks, CA: Sage Publication; 1999. https://doi.org/10.4135/9780857028075

23. World Health Organization. WHO’s Pain Relief Ladder. 2009. Available at: www.who.int/cancer/palliative/painladder/en/. Accessed November, 2017.

24. Krejcie RV, Morgan DW. Determining sample size for research activities. Educ Psychol Meas. 1970;30:607–610.

25. Altman DG. Practical statistics for medical research. London, Chapman and Hall; 1991.

26. Andrade C. Understanding relative risk, odds ratio, and related terms: as simple as it can get. J Clin Psychiatry. 2015;76(7):e857–e861.

27. Starkweather A. Persistent Pain After Breast Cancer Surgery: Risk Factors Risk Factors and Strategies to Reduce Incidence and Severity. Lippincott Williams and Wilkins. 2018;32(1):1–10.

28. Richebé P, Rivat C, Liu SS. Perioperative or postoperative nerve block for preventive analgesia: should we care about the timing of our regional anesthesia? Anesth Analg. 2013;116:969–970.

29. Hussain, et al. Should thoracic paravertebral blocks be used to prevent chronic post-surgical pain following breast cancer surgery? A systematic analysis of evidence in light of IMMPACT recommendations. Pain. 2018;159(10):1955–1971.

30. Terkawi AS, Tsang S, Sessler DI, et al. Improving analgesic efficacy and safety of thoracic paravertebral block for breast surgery: A mixed-effects meta-analysis. Pain Physician. 2015;18(5):E757–780.

31. Schnabel A, Reichl SU, Kranke P, Zahn PK. Efficacy and safety of paravertebral blocks in breast surgery: a meta-analysis of randomized controlled trials. Br J Anaesth. 2010;105(6):842–852.

32. Wahba SS, Kamal SM. Thoracic paravertebral block versus pectoral nerve block for analgesia after breast surgery. Egypt J Anaesth. 2014;30(2):129–135.

33. Kulhari S, Bharti N, Bala I, Arora S, Singh G. Efficacy of pectoral nerve block versus thoracic paravertebral block for postoperative analgesia after radical mastectomy: a randomized controlled trial. Br J Anaesth. 2016;117(3):382–386.

34. Salovey P, Smith AF, Turk DC, et al. The accuracy of memory for pain: not so bad most of the time. Am Pain Soc J. 1993;2:184–91.

35. Miranda H, Gold JE, Gore R, Punnett L. Recall of prior musculoskeletal pain. Scand J Work Environ Heal. 2006;32(4):294–300.

